# Acute heart failure patients with a high red blood cell distribution width-to-albumin ratio have an increased risk of all-cause mortality

**DOI:** 10.1101/2023.04.17.23288709

**Authors:** Shiwei Wang, Qiang Xiao, Quanqiang Lin, Yuanmin Li

**Author notes:** Correspondence: Yuanmin Li, Department of Cardiology, the Second Affiliated Hospital, Shandong First Medical University & Shandong Academy of Medical Sciences, No.366 Taian Street, Taian, 271000, P.R.China. Emaill.

## Abstract

**Background:** Many studies have shown that specific blood markers, such as red cell distribution width (RDW) and albumin levels, can provide valuable information about the prognosis of patients with acute heart failure (AHF). In light of these findings, the current study aims to investigate the relationship between another blood marker, RDW to albumin ratio (RAR), and the prognosis of AHF patients.

**Methods:** Data on patients diagnosed with AHF were extracted from the MIMIC-IV database version 2.1. Patients were divided into three groups based on RAR tertiles.

Multiple imputation was used for missing data, and pooled analysis was performed for imputed data sets. This study used Cox regression analysis to evaluate the impact of RAR on Clinical Outcomes in AHF patients. To further assess the prognostic ability of RDW, RAR, and albumin, the study also used time-dependent receiver operating characteristic (time-ROC) analysis.

**Results:** This study enrolled 1432 patients with AHF, with a mean age of 72.4 years and a mean RAR of 5.07 ± 1.51% /g/dl. Patients with AHF had increased all-cause mortality when their RAR was higher (HR = 1.16, 95% CI: 1.10 ∼ 1.23, P < 0.001), and RAR and mortality from all causes were linearly related in patients with AHF (P non-linearity = 0.643). Based on time-ROC curves, it was discovered that RAR had a higher prognostic accuracy compared to RDW and albumin.

**Conclusions:** An increased level of RAR was associated with a poor all-cause mortality prognosis for patients with AHF, and there is a significant linear relationship. RAR was a better predictor of all-cause mortality in AHF patients than RDW and albumin.

## Introduction

Heart failure (HF) continues to be a significant clinical and public health issue as the end stage of many heart diseases, with a considerable burden of morbidity and mortality^1^. The prognosis of acute heart failure (AHF) is worse than that of chronic heart failure ^2, 3^. However, some biomarkers are available to predict the prognosis of AHF patients, such as NT-proBNP^4^, troponin^5^, MR-pro-adrenomedullin^6^, Carbohydrate antigen 125^7^, sST2^8^, blood lactate^9^ and serum iron^10^, but their complexity or high price limit their widespread use. For patients with AHF, there is an urgent need for more affordable and more accessible biomarkers.

Red cell distribution width (RDW), as a parameter reflecting the degree of heterogeneity of red blood cell volume^11^, has been shown in recent years to be associated with various diseases such as cardiovascular diseases^12^, malignant diseases^13, 14^, liver diseases^15^, and autoimmune diseases^16^. It has also been shown to be a prognostic marker for AHF^17, 18^.

Albumin is a plasma protein synthesized by the liver. Serum albumin is a simple indicator that helps maintain blood volume, regulate plasma osmolality, and transport drugs^19^. It can be used to measure nutritional status and inflammatory response^20, 21^. Studies have shown that a simple way to predict the long-term prognosis of AHF patients is to measure serum albumin^22^.

Both RDW and serum albumin are commonly used clinical parameters that have the advantages of being simple, inexpensive, and easy to perform. RDW to albumin ratio (RAR), a novel biomarker, can be used to combine the predictive information of these two variables to predict the prognosis of various diseases, such as chronic heart failure, stroke, and sepsis^23-25^. To date, no studies have examined the role of RAR on the prognosis of patients hospitalized with AHF. Therefore, this study aimed to assess the predictive value of RAR in adult AHF patients.

## Methods

### Data Source

In this study, patients were chosen out of the Medical Information Mart for Intensive Care IV (MIMIC-IV) V2.1, a free, publicly available, large-sample, single-center critical care database in the United States^26^. The MIMIC-IV V2.1 database provides extensive information about 299,777 hospitalized patients during the period of 2008 to 2019. Any confidential data from this database was erased with the permission of the Institutional Review Boards of the Massachusetts Institute of Technology and Beth Israel Deaconess Medical Center. The online training course and exam (Record ID:52021956) were completed to gain access to the database.

### Population Selection Criteria

To conduct this study, we selected patients who satisfied specific conditions to participate. Selection conditions were as below: (1) The patients who have been diagnosed with AHF; (2) patients admitted to the ICU for the first time; and (3) aged ≥ 18 years. The conditions for exclusion were as below: (1) intensive care unit (ICU) stay less than 24 hours; (2) patients without RDW or albumin data; (3) patients without NT-proBNP data;

### Data Extraction

In order to extract data, we utilized Structured Query Language (SQL) and pgAdmin (version 4) software. The clinical baseline data obtained include the following parameters: ICU stay (days), age (years), gender, heart rate (beats/min), systolic blood pressure (SBP, mmHg), diastolic blood pressure (DBP, mmHg), mean blood pressure (MBP, mmHg), respiratory rate (beats/min), temperature (°C), percutaneous oxygen saturation (SPO2, %), renal replacement therapy (RRT). Additionally, comorbidities such as hypertension, diabetes, chronic obstructive pulmonary disease (COPD), and atrial fibrillation (AF) were included. Laboratory parameters such as hematocrit (%), hemoglobin (g/dl), RDW (%), platelet (10^9^/L), WBC (10^9^/L), albumin (g/dL), anion gap (AG, mmol/L), bicarbonate (mmol/L), blood urea nitrogen (BUN, mg/dL), calcium (Ca, mmol/L), chloride (mmol/L), creatinine (CR, mg/dL), glucose (GLU, mg/dL), sodium (mmol/L), potassium (mmol/L), alanine aminotransferase (ALT, IU/L), alkaline phosphatase (ALP, IU/L), aspartate aminotransferase (AST, IU/L), bilirubin total (mg/dL), and NT-proBNP (pg/mL) were also extracted. The criticality of the disease is assessed by three different scoring systems, namely the sequential organ failure assessment (SOFA) score^27^, the simplified acute physiology score II (SAPS II)^28^, and the acute physiology score III (APS III). In-hospital medications such as antibiotics, vasoactive drugs, antiplatelet drugs, β-blockers, ACEI/ARB, diuretics, nitrates, statins, and digoxin were also included. The data presented represents the first data of each indicator during ICU stay or hospitalization. The observation period started on the admission date and continued until the time of death. Obtaining death information from both hospital records and government records. All-cause mortality is defined as the endpoint.

## Statistical Analyses

Descriptive analyses were performed according to RAR tertiles divided into three groups. In this study, the continuous data that followed a normal distribution were presented as mean ± standard deviation (mean ± SD), whereas the continuous data that did not follow a normal distribution were described using median and interquartile range [M (Q1, Q3)]. The categorical variables were presented in the form of counts and percentages. To identify whether there were significant differences between the groups, statistical analyses of these variables were performed using ANOVA in the case of normal distribution, Kruskal-Wallis H-test in the case of non-normal distribution, and the chi-square test or Fisher exact test in the case of categorical variables.

To reduce the chance of false-positive or false-negative conclusions and improve data quality, the R multivariate imputation by chained equation (MICE) package was used for multiple imputation. The imputed data sets were aggregated to reach an overall conclusion^29, 30^.

The Cox proportional risk models were developed to assess the potential association of RAR with all-cause mortality in enrolled patients. The researchers also derived hazard ratios (HRs) and 95% confidence intervals (CIs) for the full analysis.

The crude model was not adjusted for any confounding variables; In model I, the covariates included sex and age; In Model II, in addition to the variables included in Model I, the researchers adjusted for ICU stay, heart rate, SBP, DBP, MBP, respiratory rate, temperature, SPO2, hypertension, diabetes, COPD, AF, hematocrit, hemoglobin, RDW, platelet, WBC, albumin, AG, bicarbonate, BUN, Ca, chloride, CR, GLU, sodium, potassium, ALT, ALP, AST, bilirubin total, NT-proBNP, SOFA, SAPS II, APS III, and RRT; In Model III, in addition to the variables included in Model II, the researchers adjusted for antibiotics, vasoactive drugs, antiplatelet drugs, β-blockers, ACEI/ARB, diuretics, nitrates, statins, and digoxin. We also converted the RAR to a categorical variable to investigate its association with all-cause mortality and derived P values for trends, aiming to compare whether the results are different when analyzed with continuous and categorical variables, respectively. Kaplan-Meier survival curves were used to analyze survival in the three RAR groups. A restricted cubic spline (RCS) model was used to observe whether there was a linear relationship between RAR and all-cause mortality in AHF patients. To investigate the existence of a linear correlation between RAR and all-cause mortality in patients diagnosed with AHF using a restricted cubic spline curve (RCS) model.

To compare the prognostic abilities of RAR, RDW, and albumin, time-dependent receiver operating characteristic (time-ROC) analysis was performed by utilizing the “timeROC” R package^31^. A P value < 0.05 was considered statistically significant.

### Subgroup Analysis

We explored whether the correlations differed between different subgroups in order to assess the robustness of the findings, including ICU stay, gender, age, hypertension, diabetes, COPD, AF, NT-proBNP, and RRT.

### Sensitivity Analysis

The following sensitivity analysis was performed to ensure the stability of the results. (1) Because NT-proBNP is an essential predictor of prognosis in acute heart failure^4^, we excluded patients without NT-proBNP data in this study, and in sensitivity analysis, we included patients without NT-proBNP data; (2) Patients who had incomplete data were excluded from the study, and a sensitivity analysis was conducted using only complete data without multiple imputation; (3) Sensitivity analysis using propensity score matching methods to balance the baseline characteristics. Using the best cut-off value of the 1-year ROC curve as the boundary, we classified the enrolled patients into two categories, the L-RAR group and the H-RAR group, based on the RAR value, and set the caliper value to 0.02. All statistical analyses were conducted using SPSS (IBM, IL, USA) version 26 and R software (Version 4.2.2, http://www.r-project.org).

## Results

### Clinical Parameters

In the MIMIC-IV V2.1, 5,694 patients with AHF and ICU admissions were among the 299,777 patients. 4,262 of these patients were not included in the study because ICU stay < 24h or missing RDW data, albumin data, or NT-proBNP data, and there were no patients < 18 years old. After screening patients using inclusion and exclusion conditions, a total of 1432 patients diagnosed with AHF were ultimately included in this study (Figure 1). A small amount of data was missing from the final 1,432 patients enrolled in this study. The missing data were as follows: respiratory rate (0.1%), temperature (0.3%), ALT (3.6%), ALP (3.6%), AST (3.5%), and bilirubin total (3.3%). Multiple imputation of the missing values was performed, and the five imputed data sets were pooled to draw overall conclusions.

**Figure 1.**
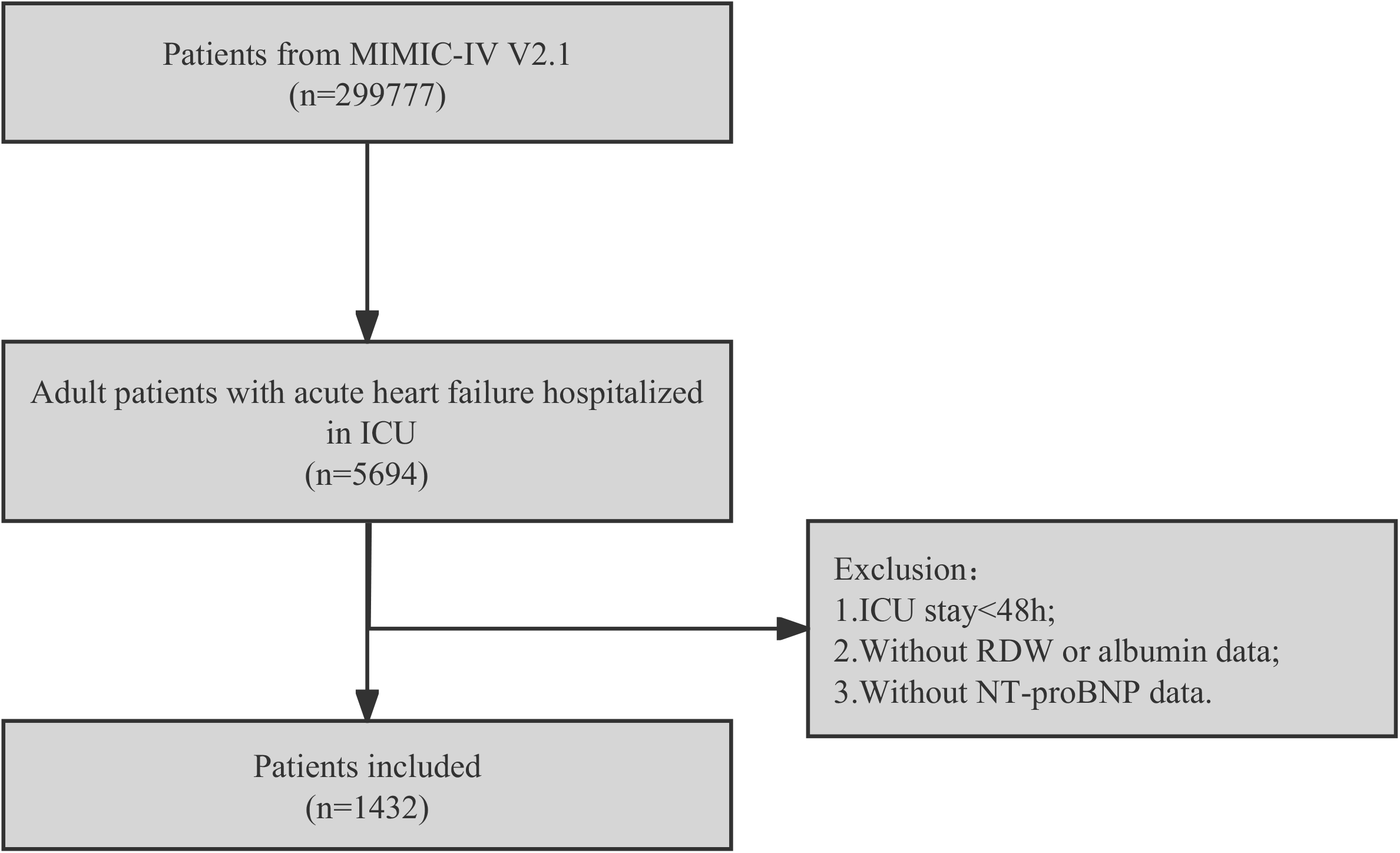
Flow chart of inclusion and exclusion. Abbreviations: MIMIC-IV: the medical information mart for intensive care IV, ICU: intensive care unit, RDW: red cell distribution width.

Figure 1 illustrates the clinical characteristics of the final enrolled patients. The enrolled patients were, on average, 72.4 years old, of whom 646 (45.1%) were female and 786 (54.9%) were male. The RAR value was, on average, 5.07 ± 1.51% /g/dl, and the ICU stay was, on average, 6.30 days. Atrial fibrillation (52.0%) and diabetes (40.0%) were the two most common comorbidities.

The enrolled patients were classified into three groups according to their RAR values, Q1 (< 4.25), Q2 (4.25 - 5.29), and Q3 (> 5.29). Increased heart rate, BUN, CR, ALP, bilirubin total, NT-proBNP, SOFA score, APS III score, and SAPS II score but decreased SBP, MBP, hematocrit, hemoglobin, AG, Ca, chloride, GLU, and ALT in patients with elevated RAR. A more significant proportion of patients with higher RAR were treated with RRT (P < 0.001) and antibiotics (P < 0.001), while a smaller proportion were treated with antiplatelet agents (P = 0.005), ACEI/ARB (P < 0.001), nitrates (P < 0.001), and statins (P = 0.009).

### Relationship Between RAR and All-cause Mortality

During a median follow-up period of 13.7 months, 739 (51.6%) patients died. As can be seen from the KM curves, a high RAR was positively correlated with a lower overall survival rate compared to a low RAR (P < 0.001, Figure 2).

**Figure 2.**
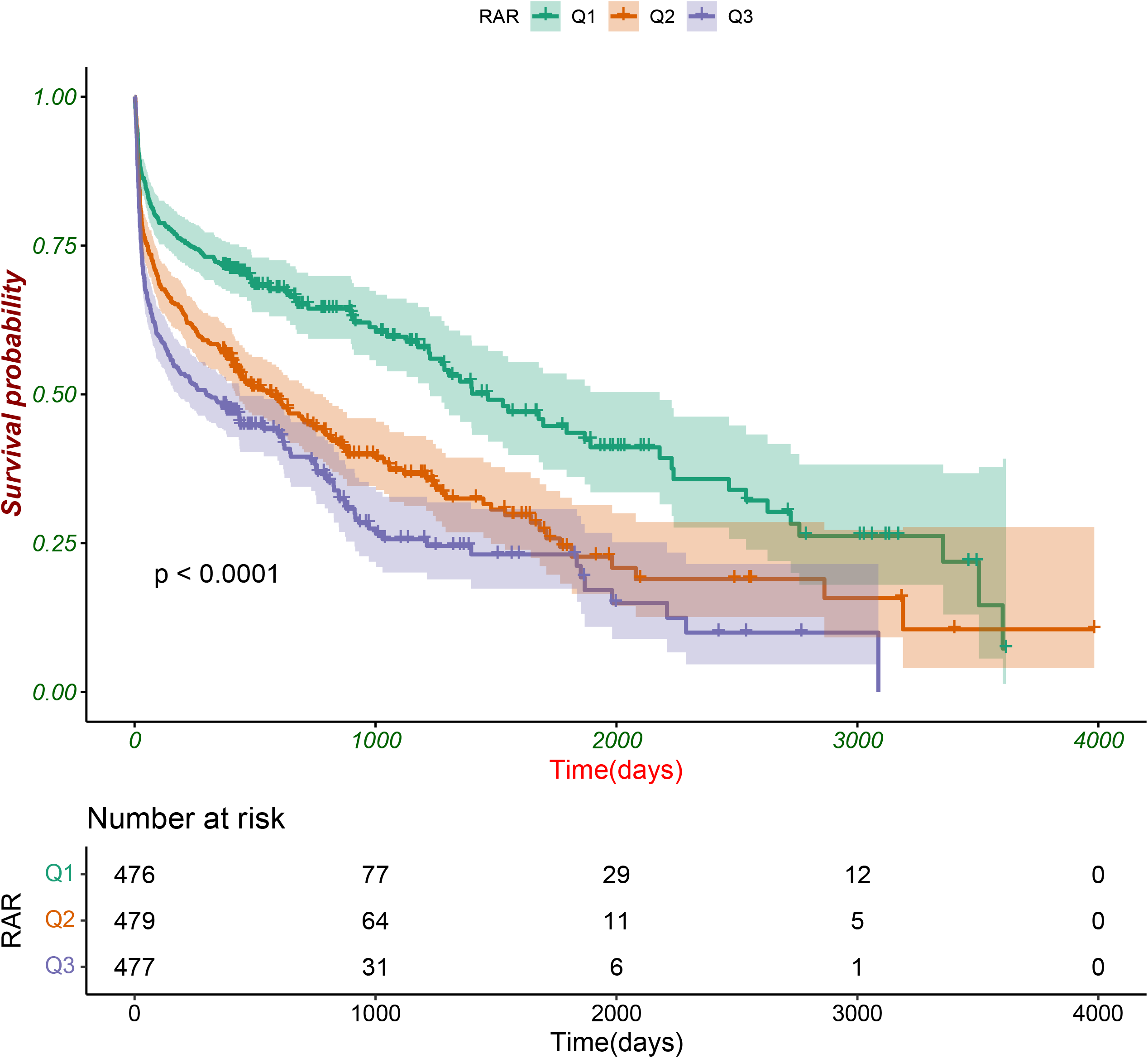
The K-M survival curve of Q1(RAR<4.25),Q2(RAR:4.25-5.29) and Q3(RAR>5.29). The range area represents a 95% confidence interval. Q: quartile, RAR: red blood cell distribution width/albumin ratio.

The COX regression analysis results relating RAR to clinical outcomes for the different models are presented in Figure 2. HRs and 95% CIs are used to describe the statistical data. Findings show that patients with elevated RAR have a higher all-cause mortality risk (Crude Model: 1.26 (1.21, 1.32), P < 0.001; Model I: 1.26 (1.21, 1.31), P < 0.001; Model II: 1.17 (1.10, 1.24), P < 0.001; Model III: 1.16 (1.10, 1.23), P < 0.001). When we converted the RAR to a categorical variable for analysis, we can see that the results for the relationship between the RAR and clinical outcomes were similar to the results as a continuous variable (Table 2). Alternatively, we found a significant trend between tertiles in several models (P for trend < 0.001, < 0.001, < 0.001, and 0.002, respectively). The association between RAR and all-cause mortality among AHF patients was examined using restricted cubic spline analysis. We found a linear relationship between RAR and all-cause mortality in AHF patients after adjusting for potential covariates (P non-linearity = 0.643, Figure 3).

**Table 1.**
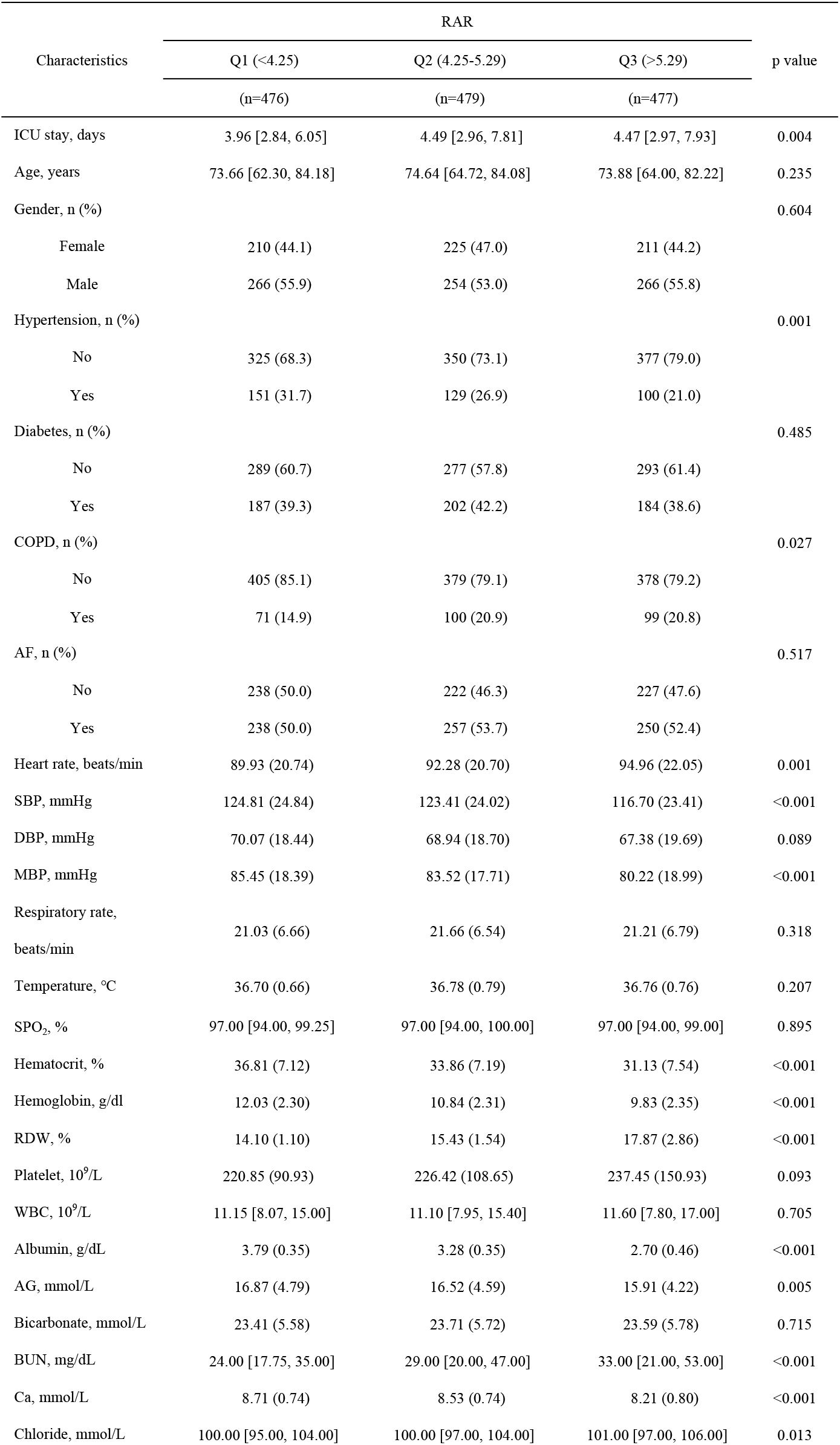
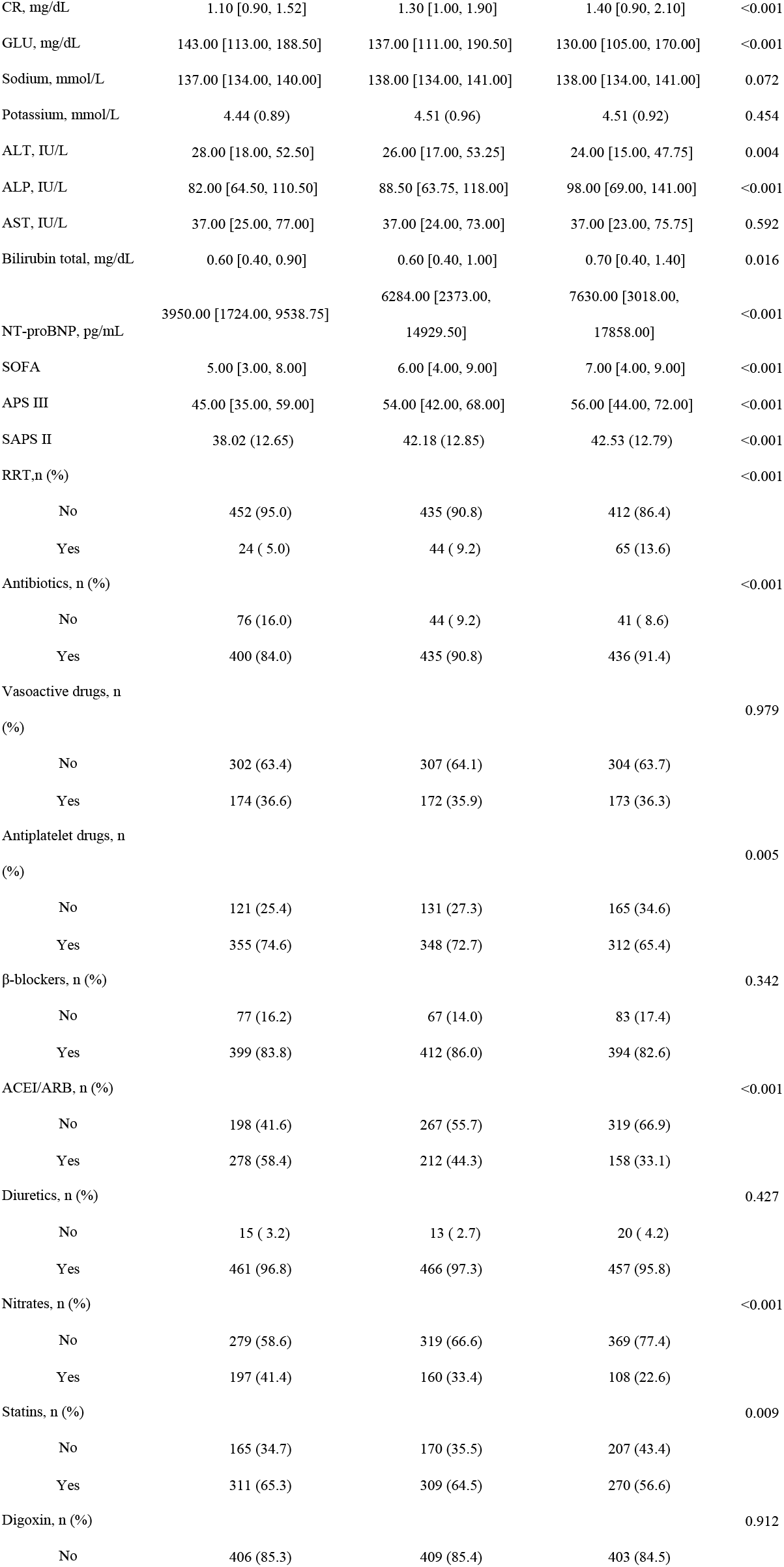
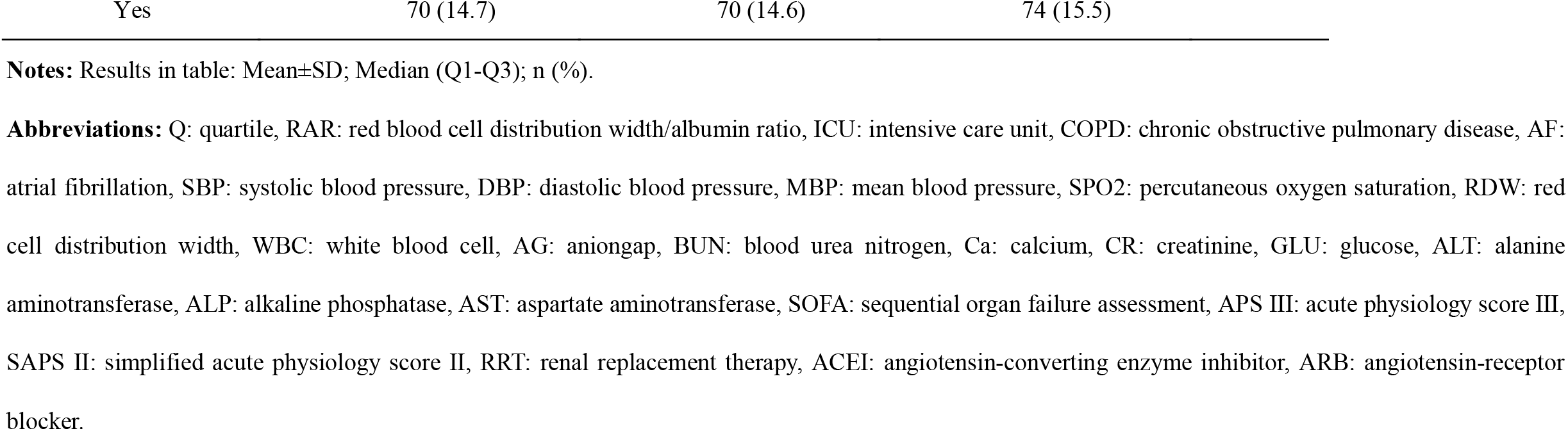
Baseline Characteristics of the Study Population.

| Characteristics | RAR |  |  | p value |
| --- | --- | --- | --- | --- |
|  | Q1 (<4.25) | Q2 (4.25-5.29) | Q3 (>5.29) |  |
|  | (n=476) | (n=479) | (n=477) |  |
| ICU stay, days | 3.96 [2.84, 6.05] | 4.49 [2.96, 7.81] | 4.47 [2.97, 7.93] | 0.004 |
| Age, years | 73.66 [62.30, 84.18] | 74.64 [64.72, 84.08] | 73.88 [64.00, 82.22] | 0.235 |
| Gender, n (%) |  |  |  | 0.604 |
| Female | 210 (44.1) | 225 (47.0) | 211 (44.2) |  |
| Male | 266 (55.9) | 254 (53.0) | 266 (55.8) |  |
| Hypertension, n (%) |  |  |  | 0.001 |
| No | 325 (68.3) | 350 (73.1) | 377 (79.0) |  |
| Yes | 151 (31.7) | 129 (26.9) | 100 (21.0) |  |
| Diabetes, n (%) |  |  |  | 0.485 |
| No | 289 (60.7) | 277 (57.8) | 293 (61.4) |  |
| Yes | 187 (39.3) | 202 (42.2) | 184 (38.6) |  |
| COPD, n (%) |  |  |  | 0.027 |
| No | 405 (85.1) | 379 (79.1) | 378 (79.2) |  |
| Yes | 71 (14.9) | 100 (20.9) | 99 (20.8) |  |
| AF, n (%) |  |  |  | 0.517 |
| No | 238 (50.0) | 222 (46.3) | 227 (47.6) |  |
| Yes | 238 (50.0) | 257 (53.7) | 250 (52.4) |  |
| Heart rate, beats/min | 89.93 (20.74) | 92.28 (20.70) | 94.96 (22.05) | 0.001 |
| SBP, mmHg | 124.81 (24.84) | 123.41 (24.02) | 116.70 (23.41) | <0.001 |
| DBP, mmHg | 70.07 (18.44) | 68.94 (18.70) | 67.38 (19.69) | 0.089 |
| MBP, mmHg | 85.45 (18.39) | 83.52 (17.71) | 80.22 (18.99) | <0.001 |
| Respiratory rate, beats/min | 21.03 (6.66) | 21.66 (6.54) | 21.21 (6.79) | 0.318 |
| Temperature, °C | 36.70 (0.66) | 36.78 (0.79) | 36.76 (0.76) | 0.207 |
| SPO<sub>2</sub>, % | 97.00 [94.00, 99.25] | 97.00 [94.00, 100.00] | 97.00 [94.00, 99.00] | 0.895 |
| Hematocrit, % | 36.81 (7.12) | 33.86 (7.19) | 31.13 (7.54) | <0.001 |
| Hemoglobin, g/dl | 12.03 (2.30) | 10.84 (2.31) | 9.83 (2.35) | <0.001 |
| RDW, % | 14.10 (1.10) | 15.43 (1.54) | 17.87 (2.86) | <0.001 |
| Platelet, 10<sup>9</sup>/L | 220.85 (90.93) | 226.42 (108.65) | 237.45 (150.93) | 0.093 |
| WBC, 10<sup>9</sup>/L | 11.15 [8.07, 15.00] | 11.10 [7.95, 15.40] | 11.60 [7.80, 17.00] | 0.705 |
| Albumin, g/dL | 3.79 (0.35) | 3.28 (0.35) | 2.70 (0.46) | <0.001 |
| AG, mmol/L | 16.87 (4.79) | 16.52 (4.59) | 15.91 (4.22) | 0.005 |
| Bicarbonate, mmol/L | 23.41 (5.58) | 23.71 (5.72) | 23.59 (5.78) | 0.715 |
| BUN, mg/dL | 24.00 [17.75, 35.00] | 29.00 [20.00, 47.00] | 33.00 [21.00, 53.00] | <0.001 |
| Ca, mmol/L | 8.71 (0.74) | 8.53 (0.74) | 8.21 (0.80) | <0.001 |
| Chloride, mmol/L | 100.00 [95.00, 104.00] | 100.00 [97.00, 104.00] | 101.00 [97.00, 106.00] | 0.013 |
| CR, mg/dL | 1.10 [0.90, 1.52] | 1.30 [1.00, 1.90] | 1.40 [0.90, 2.10] | <0.001 |
| GLU, mg/dL | 143.00 [113.00, 188.50] | 137.00 [111.00, 190.50] | 130.00 [105.00, 170.00] | <0.001 |
| Sodium, mmol/L | 137.00 [134.00, 140.00] | 138.00 [134.00, 141.00] | 138.00 [134.00, 141.00] | 0.072 |
| Potassium, mmol/L | 4.44 (0.89) | 4.51 (0.96) | 4.51 (0.92) | 0.454 |
| ALT, IU/L | 28.00 [18.00, 52.50] | 26.00 [17.00, 53.25] | 24.00 [15.00, 47.75] | 0.004 |
| ALP, IU/L | 82.00 [64.50, 110.50] | 88.50 [63.75, 118.00] | 98.00 [69.00, 141.00] | <0.001 |
| AST, IU/L | 37.00 [25.00, 77.00] | 37.00 [24.00, 73.00] | 37.00 [23.00, 75.75] | 0.592 |
| Bilirubin total, mg/dL | 0.60 [0.40, 0.90] | 0.60 [0.40, 1.00] | 0.70 [0.40, 1.40] | 0.016 |
| NT-proBNP, pg/mL | 3950.00 [1724.00, 9538.75] | 6284.00 [2373.00, 14929.50] | 7630.00 [3018.00, 17858.00] | <0.001 |
| SOFA | 5.00 [3.00, 8.00] | 6.00 [4.00, 9.00] | 7.00 [4.00, 9.00] | <0.001 |
| APS III | 45.00 [35.00, 59.00] | 54.00 [42.00, 68.00] | 56.00 [44.00, 72.00] | <0.001 |
| SAPS II | 38.02 (12.65) | 42.18 (12.85) | 42.53 (12.79) | <0.001 |
| RRT,n (%) |  |  |  | <0.001 |
| No | 452 (95.0) | 435 (90.8) | 412 (86.4) |  |
| Yes | 24 ( 5.0) | 44 ( 9.2) | 65 (13.6) |  |
| Antibiotics, n (%) |  |  |  | <0.001 |
| No | 76 (16.0) | 44 ( 9.2) | 41 ( 8.6) |  |
| Yes | 400 (84.0) | 435 (90.8) | 436 (91.4) |  |
| Vasoactive drugs, n (%) |  |  |  | 0.979 |
| No | 302 (63.4) | 307 (64.1) | 304 (63.7) |  |
| Yes | 174 (36.6) | 172 (35.9) | 173 (36.3) |  |
| Antiplatelet drugs, n (%) |  |  |  | 0.005 |
| No | 121 (25.4) | 131 (27.3) | 165 (34.6) |  |
| Yes | 355 (74.6) | 348 (72.7) | 312 (65.4) |  |
| β-blockers, n (%) |  |  |  | 0.342 |
| No | 77 (16.2) | 67 (14.0) | 83 (17.4) |  |
| Yes | 399 (83.8) | 412 (86.0) | 394 (82.6) |  |
| ACEI/ARB, n (%) |  |  |  | <0.001 |
| No | 198 (41.6) | 267 (55.7) | 319 (66.9) |  |
| Yes | 278 (58.4) | 212 (44.3) | 158 (33.1) |  |
| Diuretics, n (%) |  |  |  | 0.427 |
| No | 15 ( 3.2) | 13 ( 2.7) | 20 ( 4.2) |  |
| Yes | 461 (96.8) | 466 (97.3) | 457 (95.8) |  |
| Nitrates, n (%) |  |  |  | <0.001 |
| No | 279 (58.6) | 319 (66.6) | 369 (77.4) |  |
| Yes | 197 (41.4) | 160 (33.4) | 108 (22.6) |  |
| Statins, n (%) |  |  |  | 0.009 |
| No | 165 (34.7) | 170 (35.5) | 207 (43.4) |  |
| Yes | 311 (65.3) | 309 (64.5) | 270 (56.6) |  |
| Digoxin, n (%) |  |  |  | 0.912 |
| No | 406 (85.3) | 409 (85.4) | 403 (84.5) |  |

|  |  |  |  |
| --- | --- | --- | --- |
| Yes | 70 (14.7) | 70 (14.6) | 74 (15.5) |

**Notes:** Results in table: Mean±SD; Median (Q1-Q3); n (%).
**Abbreviations:** Q: quartile, RAR: red blood cell distribution width/albumin ratio, ICU: intensive care unit, COPD: chronic obstructive pulmonary disease, AF: atrial fibrillation, SBP: systolic blood pressure, DBP: diastolic blood pressure, MBP: mean blood pressure, SPO2: percutaneous oxygen saturation, RDW: red cell distribution width, WBC: white blood cell, AG: aniongap, BUN: blood urea nitrogen, Ca: calcium, CR: creatinine, GLU: glucose, ALT: alanine aminotransferase, ALP: alkaline phosphatase, AST: aspartate aminotransferase, SOFA: sequential organ failure assessment, APS III: acute physiology score III, SAPS II: simplified acute physiology score II, RRT: renal replacement therapy, ACEI: angiotensin-converting enzyme inhibitor, ARB: angiotensin-receptor blocker.

**Table 2.** Univariate and Multivariate Results by Cox Regression

| Variable | Crude Model (HR, 95% CI, P) | Model I (HR, 95% CI, P) | Model II (HR, 95% CI, P) | Model III (HR, 95% CI, P) |
| --- | --- | --- | --- | --- |
| **RAR** | 1.26 (1.21, 1.32) <0.001 | 1.26 (1.21, 1.31) <0.001 | 1.17 (1.10, 1.24) <0.001 | 1.16 (1.10, 1.23) <0.001 |
| **RAR (Tertiles)** |  |  |  |  |
| Q1 (<4.25) | Ref | Ref | Ref | Ref |
| Q2 (4.25-5.29) | 1.67 (1.39, 2.02) <0.001 | 1.67 (1.38, 2.01) <0.001 | 1.33 (1.09, 1.63) 0.005 | 1.34 (1.09, 1.64) 0.005 |
| Q3 (>5.29) | 2.18 (1.81, 2.63) <0.001 | 2.22 (1.84, 2.68) <0.001 | 1.54 (1.23, 1.94) <0.001 | 1.52 (1.21, 1.91) <0.001 |
| **P For Trend** | <0.001 | <0.001 | <0.001 | 0.002 |

**Notes:** Crude Model adjusted for nothing. Model I adjusted for sex, age. Model II adjusted for model I plus ICU stay, hypertension, COPD, diabetes, AF, heart rate, SBP, DBP, MBP, respiratory rate, temperature, SPO<sub>2</sub>, hematocrit, hemoglobin, platelet, WBC, AG, bicarbonate, BUN, Ca, chloride, CR, GLU, sodium, potassium, ALT, ALP, AST, Bilirubin total, NT-proBNP, SOFA, APS III, SAPS II, RRT. Model III adjusted for model II plus antibiotics, vasoactive drugs, antiplatelet drugs, β-blockers, ACEI/ARB, diuretics, nitrates, statins, digoxin.
**Abbreviations:** HR: hazard ratio, CI: confidence interval, Ref: reference, RAR: red blood cell distribution width/albumin ratio, Q: quartile.

**Figure 3.**
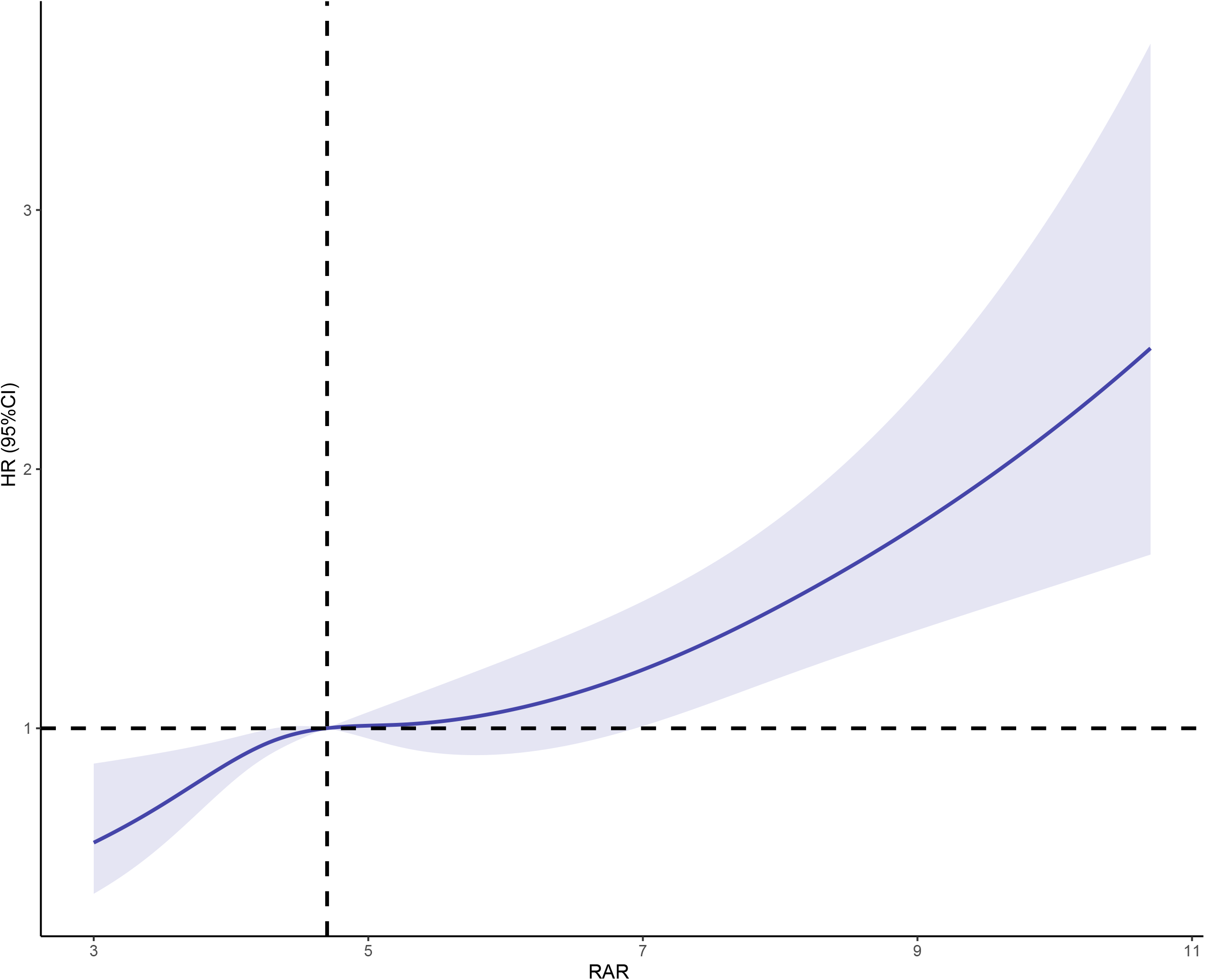
Restricted cubic spline curves of relations between RAR with all-cause mortality. There was a linear relationship between RAR and all-cause mortality, that is, the higher the RAR level, the higher the risk of death.The range area represents a 95% confidence interval. HR: hazard ratio, CI: confidence interval, RAR: red blood cell distribution width/albumin ratio.

### Receiver Operating Characteristic Analysis

We performed an analysis using time-ROC in order to further compare the predictive power of RAR, RDW, and albumin for all-cause mortality in AHF patients. Our analysis showed that RAR had a superior area under the ROC curve (AUC) (0.63, 0.63, 0.63, and 0.67) in comparison to RDW (0.59, 0.61, 0.62, and 0.63) and albumin (0.61, 0.60, 0.60, and 0.63) for predicting overall survival after 90 days, 180 days, one year, and three years (Figure 4, Figure 5). As a result, it can be said that the time-ROC curves of RAR were consistently better than those of RDW and albumin.

**Figure 4.**
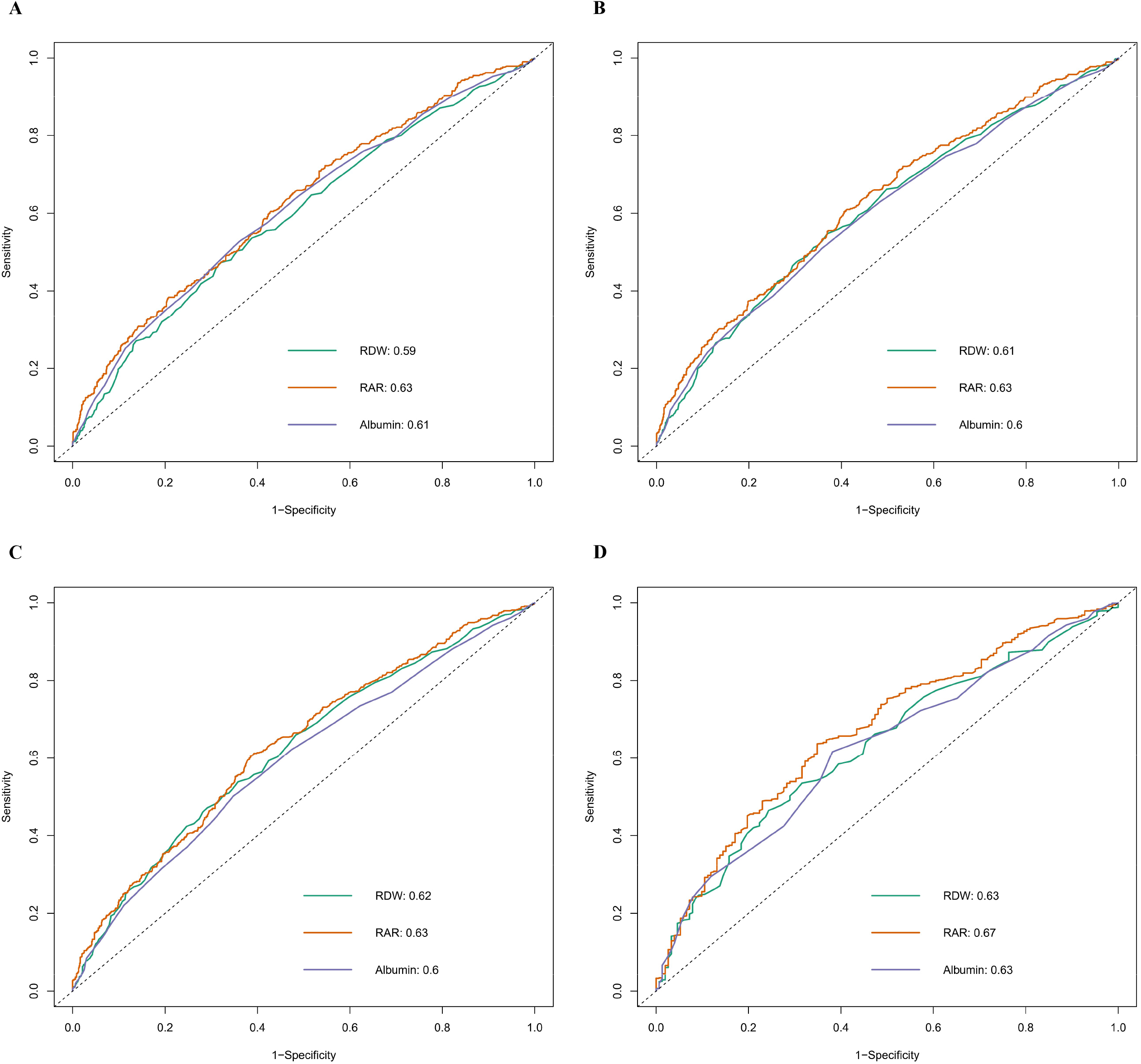
Receiver Operating Characteristic (ROC) curves of RAR, RDW and albumin for predicting all-cause mortality in AHF patients at 90 days(A), 180 days(B), 1 year(C) and 3 years(D). RDW: red cell distribution width, RAR: red blood cell distribution width/albumin ratio.

**Figure 5.**
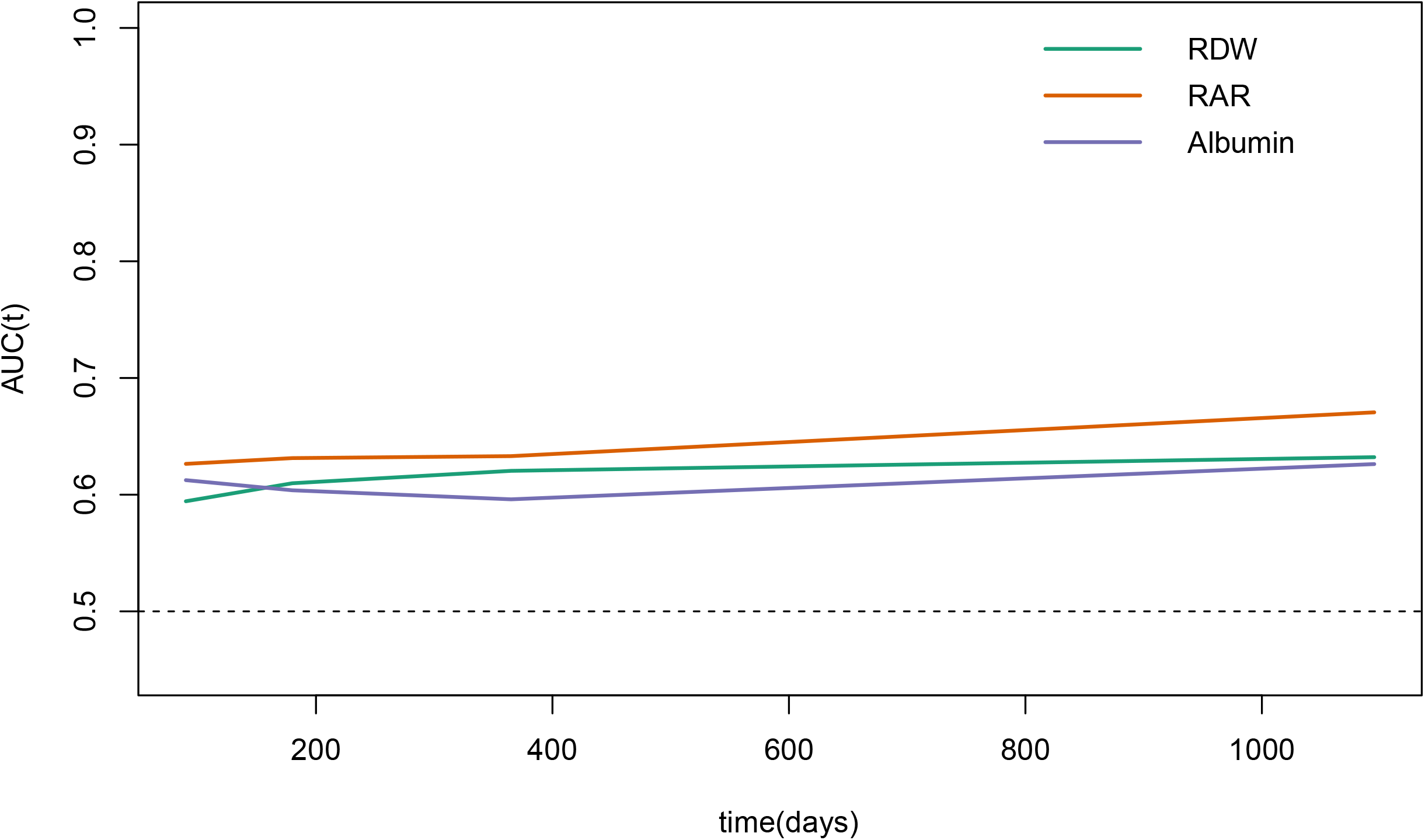
The time-ROC curve comparing the predictive value of the RAR, RDW and albumin for all-cause mortality prognosis in AHF patients. The results showed that the RAR was superior to the RDW, and albumin during follow-up. RDW: red cell distribution width, RAR: red blood cell distribution width/albumin ratio.

### Subgroup Analysis

We further investigated the effects of the additional risk between RAR and all-cause mortality in AHF patients using subgroup analysis. All tests for interaction were not statistically significant, as shown in Table 3, in subgroup analyses for ICU stay, age, sex, hypertension, diabetes, COPD, AF, NT-proBNP, and RRT (P for interaction = 0.176, 0.128, 0.399, 0.055, 0.306, 0.733 0.786, 0.960, and 0.147).

**Table 3.** Subgroup Analysis of the Relationship Between RAR Level and All-Cause Mortality

| Characteristics | N | RAR |  |  | P for interaction |
| --- | --- | --- | --- | --- | --- |
|  |  | <4.25 | 4.25-5.29 | >5.29 |  |
| **ICU stay** |  |  |  |  | 0.176 |
| <7 | 1039 | 1 | 1.20 (0.94, 1.54) 0.150 | 1.46 (1.11, 1.93) 0.008 |  |
| ≥7 | 393 | 1 | 1.71 (1.15, 5.56) 0.009 | 1.84 (1.17, 2.91) 0.009 |  |
| **Age** |  |  |  |  | 0.128 |
| <65 | 394 | 1 | 1.54 (0.91, 2.59) 0.107 | 3.09 (1.76, 5.44) <0.001 |  |
| ≥65 | 1038 | 1 | 1.32 (1.05, 1.66) 0.017 | 1.31 (1.01, 1.69) 0.046 |  |
| **Gender** |  |  |  |  | 0.399 |
| Female | 646 | 1 | 1.47 (1.07, 2.02) 0.019 | 1.64 (1.15, 2.35) 0.007 |  |
| Male | 786 | 1 | 1.23 (0.93, 1.62) 0.140 | 1.38 (1.01, 1.90) 0.045 |  |
| **Hypertension** |  |  |  |  | 0.055 |
| No | 1052 | 1 | 1.32 (1.04, 1.69) 0.023 | 1.41 (1.07, 1.85) 0.013 |  |
| Yes | 380 | 1 | 1.11 (0.74, 1.69) 0.612 | 1.59 (0.98, 2.56) 0.060 |  |
| **Diabetes** |  |  |  |  | 0.306 |
| No | 859 | 1 | 1.20 (0.91, 1.57) 0.205 | 1.55 (1.13, 2.12) 0.007 |  |
| Yes | 573 | 1 | 1.57 (1.12, 2.20) 0.009 | 1.54 (1.06, 2.22) 0.022 |  |
| **COPD** |  |  |  |  | 0.733 |
| No | 1162 | 1 | 1.34 (1.07, 1.68) 0.011 | 1.62 (1.25, 2.09) <0.001 |  |
| Yes | 280 | 1 | 1.43 (0.86, 2.38) 0.173 | 1.21 (0.69, 2.13) 0.505 |  |
| **AF** |  |  |  |  | 0.786 |
| No | 687 | 1 | 1.47 (1.08, 2.00) 0.016 | 1.87 (1.32, 2.66) <0.001 |  |
| Yes | 745 | 1 | 1.27 (0.95, 1.69) 0.110 | 1.36 (0.99, 1.88) 0.058 |  |
| **NT-proBNP** |  |  |  |  | 0.96 |
| <4000 | 579 | 1 | 1.28 (0.89, 1.84) 0.189 | 1.51 (1.01, 2.26) 0.047 |  |
| ≥4000 | 853 | 1 | 1.32 (1.02, 1.71) 0.034 | 1.54 (1.15, 2.05) 0.004 |  |
| **RRT** |  |  |  |  | 0.147 |
| No | 1299 | 1 | 1.36 (1.10, 1.69) 0.005 | 1.52 (1.19, 1.95) 0.001 |  |
| Yes | 133 | 1 | 0.83 (0.32, 2.15) 0.693 | 1.67 (0.66, 4.21) 0.280 |  |

**Notes:** HRs (95% CIs) were derived from Cox proportional hazards regression models. Covariates were adjusted as in model III (Table 2).
**Abbreviations:** RAR: red blood cell distribution width/albumin ratio, ICU: intensive care unit, COPD: chronic obstructive pulmonary disease, AF: atrial fibrillation, RRT: renal replacement therapy.

### Sensitivity Analysis

For the purpose of evaluating the study’s conclusions’ robustness, three sensitivity analyses were performed. Firstly, patients without NT-proBNP data were included, but it was excluded from Models II and III due to excessive missing values of NT-proBNP. The conducted analyses indicated that the correlation between RAR and all-cause mortality in AHF patients was still robust and significant (Supplemental Table 1). Throughout the observation period, the time-ROC curves of RAR were consistently better than those of RDW and albumin (Supplemental Figure 1). Secondly, we removed patients with missing values and performed sensitivity analysis again, with similar results as before (Supplemental Table 2, Supplemental Figure 2). Finally, we performed sensitivity analysis by applying the method of propensity score matching. We classified the patients into two categories, the L-RAR group (<4.76) and the H-RAR group (>4.76), using the best cutoff value of 4.76 for the 1-year ROC curve as the boundary. Supplemental Figure 3 shows the distribution of propensity scores. Covariates that were not balanced between the two groups were balanced after matching (p>0.05, Supplemental Table 3). The findings were similar to those of our previous study (as shown in Supplementary Table 4), confirming that RAR continues to show a significant correlation with all-cause mortality in AHF patients. The time-ROC curves showed that the AUC of RAR was slightly better than those of RDW and albumin (Supplemental Figure 4).

## Discussion

As far as we know, it is the first study to investigate the relationship between RAR and prognosis in AHF patients. In this study, the increased risk of all-cause mortality was positively and linearly associated with an elevated RAR after several adjustments for potential confounders. In comparison to RDW and albumin, a biomarker called RAR may provide better prognostic information for predicting prognosis in AHF patients.

RDW is widely used in clinical practice to reflect the degree of heterogeneity of erythrocyte volume, which can be obtained from blood cell analysis and has the advantage of being simple and inexpensive^11^. Red cell volume standard deviation (SD) divided by mean corpuscular volume (MCV) gives normal values for RDW between 11% and 15%^32^. Many studies have shown that higher RDW independently predicts poor outcomes in AHF patients^18, 33^. Pascual-Figal et al. found that RDW was a strong and independent long-term prognostic marker regardless of the anemia status of AHF patients^17^. The exact reason why RDW is associated with the prognosis of AHF patients is unknown but may be related to mechanisms such as inflammation, oxidative damage, malnutrition, and hypoxia^34-38^.

Serum albumin is the most abundant and versatile protein in human blood, accounting for 50% of plasma protein. It plays a crucial role in maintaining blood volume, regulating plasma osmolality, and transporting drugs^19^. Serum albumin has been widely demonstrated to be a strong prognostic indicator in cardiovascular disease by many studies^22, 39^. The mechanisms of the anti-inflammatory, antioxidant, anti-coagulant, and anti-aggregatory capacities of serum albumin may underlie the pathophysiology of albuminemia that affects the prognosis of cardiovascular disease^40^.

The ratio of RDW to albumin, known as RAR, is a novel biomarker that combines these two parameters to provide a more accurate prediction. Over the past few years, this biomarker has gained a lot of attention and has been extensively studied in various diseases. Patients with heart failure showed a worse prognosis when RAR was elevated, according to Ni et al.^23^. Xu et al. reported that RAR was substantially linked to a bad clinical prognosis in sepsis^24^. In a study by Zhao et al., RAR can be used to predict stroke-associated infections and mortality in stroke patients^25^. To date, the relationship between RAR and the prognosis of AHF patients has not been studied.

In AHF patients, RAR is an effective predictor of all-cause mortality and outperforms RDW and albumin, as evidenced by the AUC. RAR can be measured in the lab quickly, simply, and affordably and is not dependent on unstable variables like body temperature, heart rate, and respiratory rate. As a result, RAR has emerged as a simple yet relatively dependable biomarker that can aid in identifying patients at high risk of AHF and provide assistance in the diagnosis and treatment of AHF, especially in some less developed areas.

Because NT-proBNP is an essential predictor of prognosis in acute heart failure^4^, patients with missing NT-proBNP were excluded from this study. We incorporated patients with missing NT-proBNP in the sensitivity analysis to ensure the reliability of the study results and found that there was still a strong association between RAR and clinical outcomes in patients with AHF.

However, there are some shortcomings in this study. First, the data utilized in this study were sourced from the MIMIC-IV, a retrospective study conducted at a single center, it is possible that potential bias may exist. Therefore, it is imperative to conduct further multicenter studies to validate our findings. Secondly, it is worth noting that some variables were not incorporated into the study’s analysis due to a high number of missing values. This exclusion may have led to an imperfect model, and thus, it is important to acknowledge this limitation when interpreting the results. Third, since this study is observational, we cannot rule out the possibility that unmeasured variables had an impact on the findings. To verify our results, further studies are required. Despite these limitations, the predictive power of RAR for the prognosis of AHF patients is undeniable.

## Conclusion

The research findings suggest that an increased level of RAR was associated with a poor all-cause mortality prognosis for patients with AHF and that there is a significant linear relationship. RAR had a superior predictive ability in comparison to RDW and albumin for predicting overall survival. Therefore, RAR has the potential to become a valuable new biomarker for patients with AHF.

## Data Availability

The MIMIC-IV V2.1 was used in this study. The database can be downloaded here: https://physionet.org/content/mimiciv/2.1/.

